# Co-developing a comprehensive disease policy model with stakeholders: the case of malaria during pregnancy

**DOI:** 10.1101/2024.09.10.24313438

**Authors:** Silke Fernandes, Andrew Briggs, Kara Hanson

## Abstract

**Introduction:** Understanding the holistic impact of malaria during pregnancy is essential for improving maternal and child outcomes in malaria endemic settings. To be able to design appropriate research and conduct robust policy analyses, a comprehensive model of the underlying disease, representing the current understanding of mechanisms and consequences is essential. This study aimed to illustrate a methodology to co-develop a disease model with expert stakeholders using malaria during pregnancy as a case study.

**Methods:** An initial steering group was convened to develop a first model of malaria during pregnancy and its consequences for mother and child based on their understanding of the literature. Subsequently, this model was refined using a Delphi process to gain consensus amongst twelve experts, representing the disciplines of health economics, mathematical modelling, epidemiology and clinical medicine, working in the field of malaria during pregnancy. Experts reviewed drafts of the conceptual model and provided feedback in two rounds of semi-structured questionnaires with the aim of identifying the most important health outcomes and relationships in both mother and child as well as the most relevant stratifiers for the model. Consensus on any final disagreement was reached after two consensus meetings.

**Results:** The final model is a comprehensive disease model of malaria during pregnancy, including ten maternal and ten child outcomes with four stratifiers. The model developed in this study should be of value to malaria researchers, funders, evaluators and decision makers, though some adaptation will be required for each specific context and purpose. In addition, the methodology and process followed in this study is replicable and can guide researchers aiming to develop a conceptual model for other conditions.

**Discussion & Conclusion:** The model resulting from this study highlights the complexity required to depict appropriately the consequences of malaria during pregnancy for both the mother and the child. It also demonstrates how to conduct a rigorous process to develop a disease model. In addition the study has helped to identify a number of areas with scarce data and need for further research.

**Funding:** This study from part of the IMPROVE and IMPROVE-2 studies, which received financial support from the EDCTP2 programme under Horizon 2020 (TRIA.2015-1076, TRIA.2015-1076b); the UK Department of Health and Social Care, the UK Foreign Commonwealth and Development Office, the UK Medical Research Council, and Wellcome Trust, through the Joint Global Health Trials scheme (MR/P006922/1); and the Swedish International Development Cooperation Agency.

## Introduction

Most diseases work via complex biological processes that create observable interrelated health outcomes. Understanding these relationships is essential for many types of research and the embedded policy analyses that are informed by that research. For example, trials of comparative effectiveness and associated cost-effectiveness analysis would benefit from an understanding of all the relevant outcomes, and the interconnections among them, both in designing the most appropriate research study and in analysing the results of that study to understand which treatment options are most appropriate in a given context.

Cost effectiveness analyses (CEA) are required by many global and national bodies to inform policy change. Some health interventions have multiple health effects (for example, both morbidity and mortality); and some have effects on more than one population group. To capture such disparate benefits in a way that facilitates comparison between alternative uses of scare resources, health benefits of interventions are translated into a common metric such as a Quality Adjusted Life Year (QALY) or Disability Adjusted Life Year (DALY).

Complexity is increased when a disease such as malaria is combined with pregnancy, as the disease and its treatments now have the potential to impact both mother and child. In this paper, we use malaria in pregnancy as a case study to illustrate how a structured approach to co-developing a disease model with relevant stakeholders can result in a more robust model, that will carry greater influence with the scientific community because of the multi-disciplinary input into its development.

In 2012 a taskforce recommended the development of a conceptual model as the foundation for developing an economic model (1). A conceptual model entails a systematic approach to provide a visual framework for analysis that shows how specific health outcomes and pathways relate and interact with each other (1). Documented approaches to the development of conceptual frameworks include literature reviews, consultation with stakeholders (qualitative and quantitative), methods of incorporating stakeholder views and piloting to refine the framework (2, 3).

Economic evaluations of interventions can be complex and require contributions from a broad range of disciplines. Models based on a particular viewpoint can lead to poor validity and credibility. A review of outcomes included in published economic models of malaria in pregnancy interventions reveals a disparate picture of outcomes included in DALY estimation (4–12). Only four out of nine CEAs incorporated clinical malaria, maternal anaemia and low birth weight (5, 6, 9, 11), which are commonly measured in clinical trials, in the DALY estimation. One CEA did not include any child health outcome (8) and another no maternal health outcome (4).

The Delphi consultation method is a well-established and tested approach used in research to elicit information from experts and has been used extensively in the social sciences (13–16). It is a particularly suitable method to incorporate a range of stakeholder views, leading to improved quality and acceptance of an economic evaluation model and its findings(17) (1, 18, 19) by seeking consensus amongst experts, avoiding the pitfall of only including outcomes and relationships measured in trials.

The aim of this study was to co-develop a conceptual model of prevention of *P.falciparum* malaria during pregnancy for pregnant women and their babies using a Delphi consensus study with experts in the field of malaria during pregnancy. The expert panel’s task was to identify the most important health outcomes and relationships in both mother and child and the most relevant stratifiers for the model. In doing so, the study demonstrates that co-production of holistic disease models with expert stakeholders, representing the current understanding of a disease and potential treatment pathways, is feasible and represents a more robust approach than ad hoc model construction by individual academic teams.

## Methods

This study used the Delphi methodology to co-develop a policy model of malaria during pregnancy with expert stakeholders. The expert panel in the Delphi methodology consists of people with relevant insight into the subject to be explored and can include technical experts, health providers, policy makers, patients or other suitable panellists. It is a very useful technique to gather input from various stakeholders in a time-efficient manner through a series of questionnaires. Responses from each round are collated, analysed and incorporated into the subsequent rounds of questions until consensus between the panellists has been reached, usually after two-to-three rounds, which is often followed by a final consensus meeting with stakeholders to resolve any final points (15, 16, 20). The experts remain anonymous in the process up until the final meeting if applicable, which promotes equal contribution independent of status and other factors and removes less favourable forces of group dynamics (15, 16). The

The different stages and methods in this study are illustrated in figure 1 and summarized below. Full details of the approach are described in the supplementary materials. Experts in this study were first approached on 31 August 2022, all experts had consented by 5^th^ of October 2022. The final consensus meeting took place on 8^th^ September 2023.

**Figure 1:**
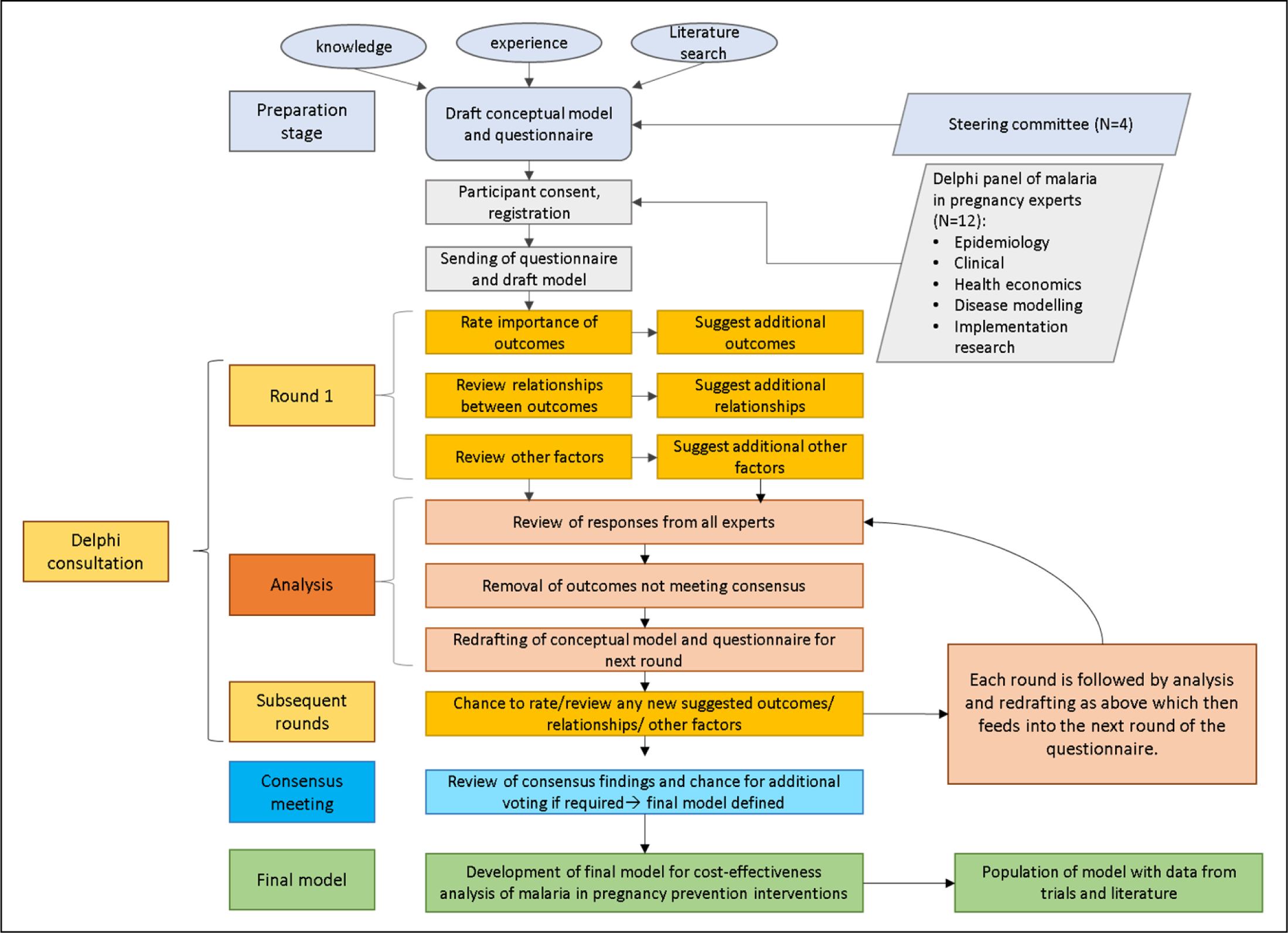
Methodology used in the study. illustrates the methodology used in this Delphi consultation study, which can be split into three stages: preparation, Delphi consultation and consensus meeting.

### Stage one: preparation

During an initial preparation stage, a steering group with collective experience in health economics, conceptual modelling and epidemiology of malaria in pregnancy was convened. Its task was to short-list experts to be approached to be part of a Delphi panel as well as to advise on the preparation of a first draft of the conceptual model and questionnaire based on their understanding of the literature, ongoing research and natural history of malaria in pregnancy. Potential candidates for the Delphi panel were purposively selected for their varied expertise, knowledge of the literature and current research in malaria during pregnancy and approached by email. The authors paid particular attention to having a well-balanced panel with experts representing both maternal and child health, early and later exposure to malaria during pregnancy and various endemicity contexts. The study team aimed to include eight to ten experts in the Delphi panel, a group size shown to be effective and reliable for the Delphi method (15, 16). Experts received no incentive or financial reimbursement for their time participating in this study.

### Stage two: Delphi consultation

Twelve expert agreed to take part in the Delphi study and provided written informed consent (online). During the Delphi consultation stage, they were asked to refine the draft model using an *a priori* undetermined number of rounds of consultations until consensus in most questions was reached. The threshold for consensus for individual questions was set at 70%, consistent with previous Delphi studies (18, 21). In each round, the panel members were provided with a current draft of the model and were asked to perform the following tasks:

1. Assess importance of the outcomes in the model,
2. suggest additional outcomes that were missing,
3. evaluate the accuracy of relationships between outcomes,
4. suggest additional relationships between outcomes that were missing,
5. order stratifiers for subgroup analysis by importance (a stratifier is defined as a variable which can partition the population in the model into subpopulations, e.g. by gravidity or HIV status of the mother),
6. suggest any additional stratifiers that were missing, and
7. provide their opinion on additional aspects of the presentation of the model, e.g. the visual presentation of low birth weight with its sub-categories (prematurity, intrauterine growth restriction and small for gestational age).

Nominal and ordinal categorical response options as well as free-text questions were used in both questionnaires with round two containing more of the latter (See appendix 2 and 3 for questionnaires used in round 1 and 2). The responses to each round were analysed by one researcher (SF) and incorporated into the next model draft and questionnaire. Categorical questions were analysed using simple descriptive statistics. Free text responses were explored using a simple thematic analysis, coding them manually into themes (22). After each round panellists received a summary report of the analysis (See appendix 4 and 5), ensuring anonymity was maintained.

### Stage three: Consensus meeting

In the final stage of the study, two online consensus meetings for experts in different time zones were held to present the findings of the second Delphi round and to discuss and vote on any remaining aspects of the model where consensus had not been reached during stage two. The conceptual model was finalized by the first author following the consensus meeting.

### Ethics

Ethics approval for this study was received on 12 July 2022 by the Research Ethics Committee of the London School of Hygiene and Tropical Medicine (Reference number 27361). Informed written consent was received from all Delphi panel members.

## Results

Key results of the different stages of this study are summarized here (see supplementary materials for additional detail).

### Stage one: Preparation

The first draft of the conceptual model developed by the steering group included outcomes for the mother and the child and relationships among them (Figure 2). Gravidity, timing of exposure to *P.falciparum* (i.e. first, second or third trimester) and HIV status were selected as the most relevant stratifiers for subpopulation analysis. The steering group identified 17 experts to be approached to participate in the study, of whom twelve agreed (71%). Amongst eleven experts the average years of experience working in malaria in pregnancy was 17.9 years (range 8-34) and the twelfth expert had over 15 years of experience in the economics of malaria.

**Figure 2:**
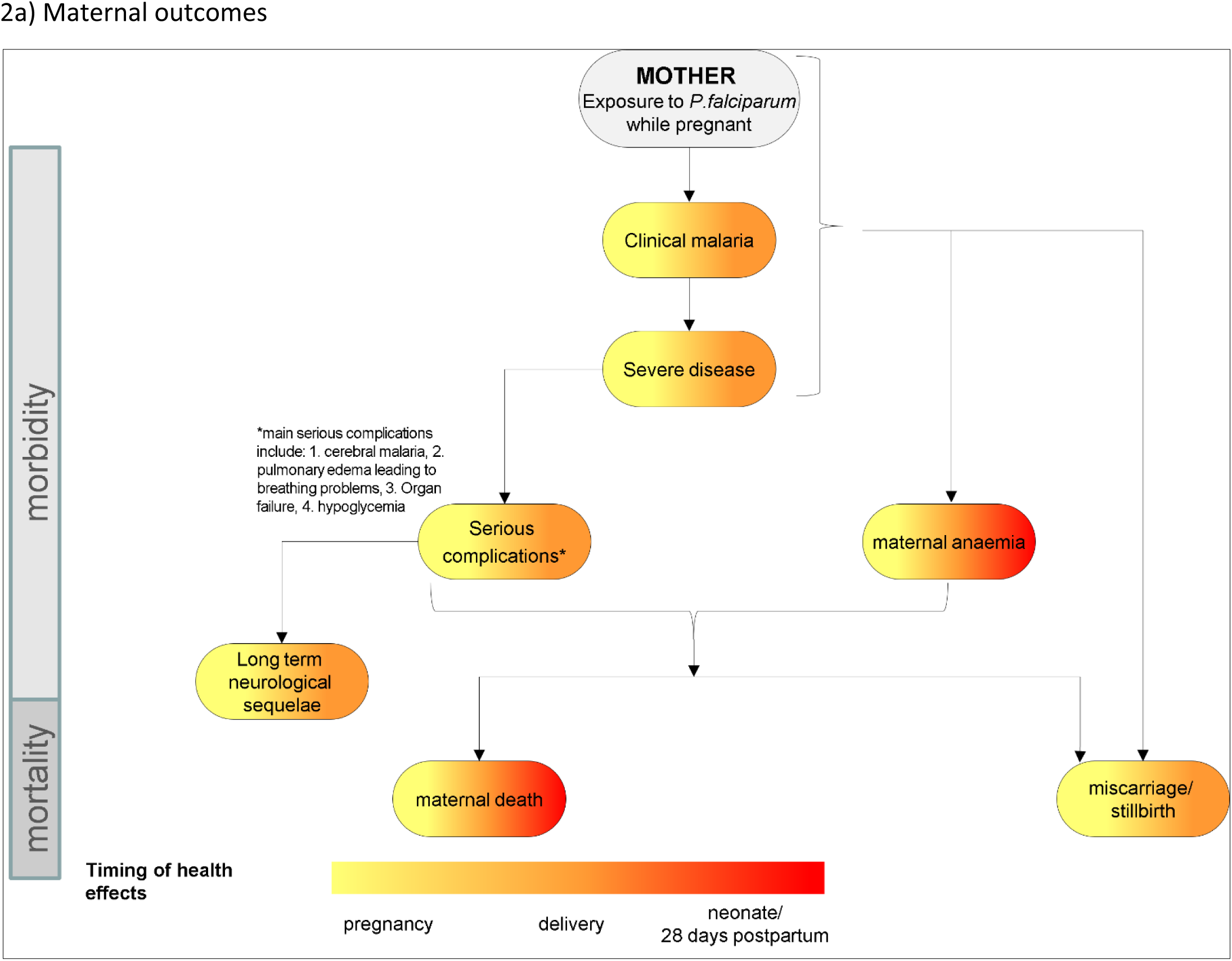
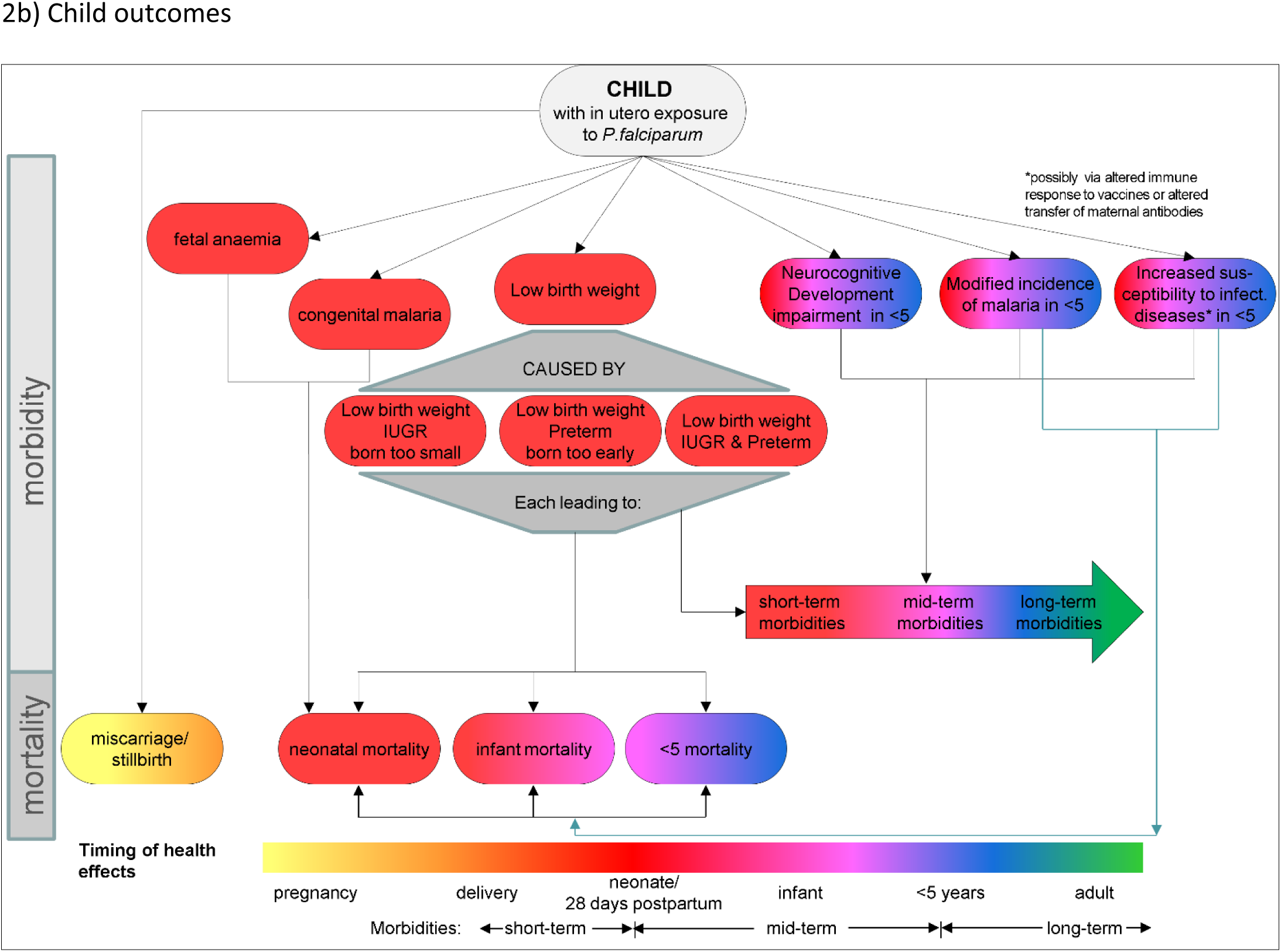
Conceptual model: Draft 1- steering group. shows the maternal and child outcomes included in the first draft of the conceptual model developed by the four members of the steering group to be used as a starting point for the first round of the Delphi consultation. Outcomes were divided into maternal and child outcomes. Outcomes affecting morbidity are shown at the top, while mortality outcomes at the bottom. The colour coding of the outcomes represents the different timings of the health effects. Abbreviations: IUGR=Intrauterine growth restriction

### Stage two: Delphi consultation

Two rounds of consultation were required before sufficient consensus was reached. Changes to the model made after each consultation round and the consultation meetings are illustrated in table 1 with only the most significant changes highlighted here.

**Table 1:** Changes to the model made after each consultation round and consultation meetings.

|  |  | Changes made after Delphi consultation round 1 | Changes made after Delphi consultation round 2 | Changes made after Delphi consultation meetings (final model) |
| --- | --- | --- | --- | --- |
| Mother | Outcomes | "MOTHER Exposure to P.falciparum while pregnant" changed to "MOTHER presence of P.falciparum while pregnant" |  |  |
|  |  | Addition of "asymptomatic parasitaemia" |  |  |
|  |  | "With and without Placental malaria" added to "clinical malaria" and "asymptomatic parasitaemia" |  |  |
|  |  | "Long term neurological sequelae" changed to "long term sequelae" to include other long-term disabilities resulting from manifestations of severe malaria | "Long term sequelae" changed to long term neurological and other sequelae" | green colour (for long-term consequence to the mother) added to "long-term neurological and other sequelae" |
|  |  | "Death in utero after maternal death" added to "miscarriage/stillbirth" | "miscarriage/stillbirth/death in utero after maternal death" changed to "miscarriage/stillbirth/death in utero" |  |
|  |  | "Hypertensions disorders of pregnancy" added | Outcome "hypertension disorders of pregnancy" relabelled to "hypertension disorders of pregnancy and post-partum", footnote added: "includes pre-eclampsia, eclampsia and gestational hypertension" | Additional outcome "Long-term effects of hypertension disorders of pregnancy" added as a consequence of "Hypertension disorders or pregnancy and post-partum" |
|  |  | "Severe disease" and "serious complications" combined into "severe malaria" and WHO definition added |  |  |
|  | Relationships | Relationships to and from "hypertension disorders of pregnancy" added |  | Relationship from "Hypertension disorders of pregnancy and post-partum" to "Long-term effects of hypertension disorders of pregnancy" added |
|  |  | Relationship from maternal death to "death in utero after maternal death" added |  |  |
|  |  | Relationship to and from "asymptomatic parasitaemia" added |  |  |
|  |  | Relationship from maternal anaemia to "severe malaria" added |  |  |
|  |  | Relationship between maternal anaemia and clinical malaria made bi-directional | Bidirectional relationship between "clinical malaria" and maternal anaemia" reversed to unidirectional (malaria to anaemia) and an arrow from "maternal anaemia" to "clinical malaria" indicating "contributes to progression" | Arrow from "maternal anaemia" to "clinical malaria" indicating "contributes to progression" removed |
| Child | Outcomes | "Small for gestational age" added and visualization of "low birth weight" changed |  |  |
|  |  | "short-, mid- and long-term morbidities" changed to "neonatal, infant, <5 and older child/adult morbidities" | "Neonatal, infant, <5 and older child/adult morbidities" changed to "Other morbidities in neonates, infants, <5, older children and adults" | Shape of "Other morbidities in neonates, infants, <5, older children and adults" changed from large arrow to an oval shape as other outcomes |
|  |  | "CHILD with in utero exposure to P.falciparum" changed to "CHILD Presence of P.falciparum in utero" | "CHILD Presence of P.falciparum in utero" relabelled to "CHILD Presence of/ exposure to P.falciparum in utero" |  |
|  |  |  | "Neonatal, infant and <5 mortality" combined into 1 large outcome box and relabelled as "Neonatal, infant, <5 mortality & mortality in older children & adults" | green colour added to box "Neonatal, infant, <5 mortality & mortality in older children & adults" |
|  |  |  | "Modified incidence of malaria in <5" changed to "Increased incidence of malaria in <5" |  |
|  |  |  |  | "Neurocognitive development impairment in <5" relabelled to "Neurocognitive & physical development impairment in <5" |
|  | Relationship | Arrow from "neonatal, infant, <5 and older child/adult morbidities" to "neonatal, infant and <5 mortality" added | Starting position of arrow from "neonatal, infant, <5 and older child/adult morbidities" to "neonatal, infant and <5 mortality" changed from back of the box to the middle |  |
|  |  |  |  | Arrows from<br>1) "CHILD Presence of/exposure to P.falciparum in utero", 2)"Fetal anaemia", 3) "Congenital malaria" to "Other morbidities in neonates, infants, <5, older children and adults" added |
| Other | Design | Symbols in addition of colour code for the timing of health effects added |  |  |
|  |  | Red on timeline changed from "neonate/ 28 days postpartum" to "neonate/mother 28/42 days postpartum" to reflect the postpartum period in which maternal death are counted | description box moved below model |  |
|  |  | Design of certain arrows and lines changed to help with distinguishing them | Design of arrows and lines changed again as previous change was confusing to expert |  |
|  | Stratifiers | "Transmission intensity" added as a stratifier to the next round in addition to "HIV status", "Gravidity" and "Timing of exposure" | Out of the four stratifiers "Transmission intensity", "HIV status", "Gravidity" and "Timing of exposure", Transmission intensity and gravidity were considered most important by experts. | Description box listing potential other stratifiers added below the model |

After the first consultation round all outcomes included in figure 2 remained in the model. On recommendation of panel members “severe disease” and “serious complications” were combined into a single outcome of “severe malaria” as experts pointed out the difficulty in differentiating between these two outcomes. All experts agreed that “low birth weight” should be separated into “intrauterine growth restriction” and “preterm birth”, with five experts suggesting the addition of “small for gestational age”. Additional outcomes - all maternal - to be incorporated into the next draft of the model were “asymptomatic parasitaemia”, “placental malaria” and “hypertension disorders of pregnancy”. Responses on rating stratifiers (gravidity, HIV status, timing of exposure) needed further exploration with transmission intensity suggested as an additional stratifier by seven experts.

In round two experts reached consensus regarding the inclusion of asymptomatic parasitaemia (100%, 12/12 agreed) and placental malaria (75%, 9/12 agreed) and their associated relationships. Whether to include or exclude “hypertension disorder of pregnancy” was unclear and had to be explored further during the consensus meeting. While all relationships associated with “hypertension disorder of pregnancy” were judged to be correct, it appeared that there was a difference in opinion regarding its importance and relevance amongst experts working in low-versus high endemicity settings.

Experts were asked to vote for the two most important stratifiers for subpopulation analysis leading to the following ranking from most to least important with the number of votes in brackets: gravidity (10), transmission intensity (8), timing of exposure of P.falciparum (3) and HIV status (2). Summary reports of both Delphi consultation round analyses can be found in appendix 4 (round 1) and 5 (round 2) and intermediate model drafts after round 1 and 2 are depicted in figure S1 and S2 in appendix 1.

### Stage three: consensus meeting

All twelve experts completed both rounds of questionnaires and nine (75%) attended one of the consensus meetings, held on 31^st^ August and 8^th^ of September 2023. The most relevant topic discussed was “hypertension disorders of pregnancy” and its potential sequelae. All attending experts agreed to keep “hypertension disorder of pregnancy” in the model without splitting it further into “hypertension”, “pre-eclampsia” and “eclampsia”. However, they voted to add “long-term effects of hypertension disorders of pregnancy” as a further outcome to include long-term sequelae such as stroke or mental health disorders. Other less contentious issues such as the relationship between “clinical malaria” and “anaemia” or relationships and labelling of child morbidities were also agreed during the consensus meeting.

Experts expressed the importance of adapting economic models to context and allowing the flexibility for them to evolve over time as more granular data become available. They also felt that in addition to developing a conceptual model of malaria during pregnancy to be used in future cost-effectiveness analysis, the work had helped to identify a number of areas where data are limited and that it will be important to share these with the research community. The final model is shown in figure 3, in which both child and maternal figures are combined, a suggestion made during the consensus meeting.

**Figure 3.**
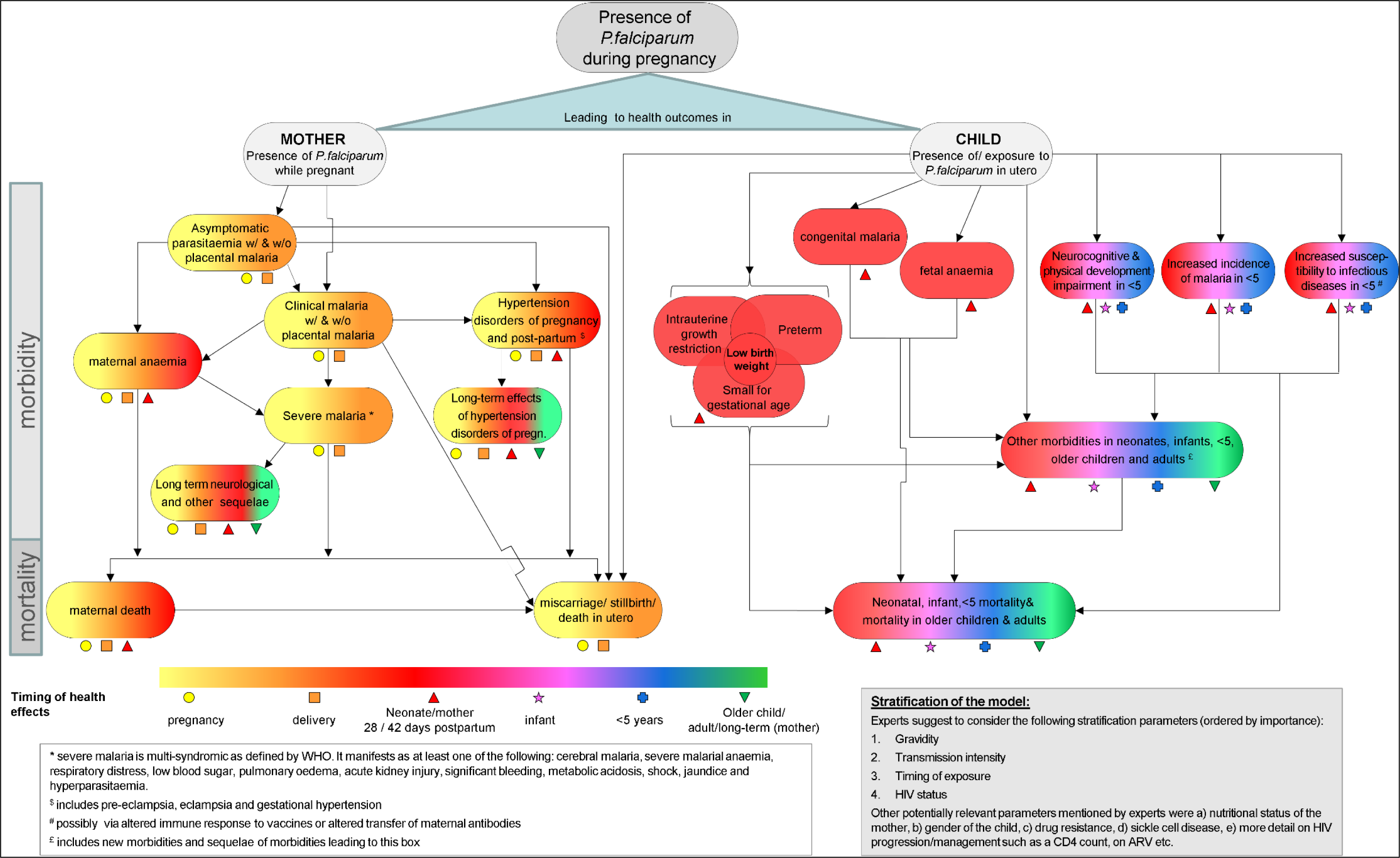
Final model after Delphi consultation meetings. The final agreed conceptual model of malaria during pregnancy following two consensus meetings attended by nine experts. The model combines both maternal and child outcomes. Outcomes were divided into maternal and child outcomes. Outcomes affecting morbidity are shown at the top, while mortality outcomes at the bottom. The colour and shape coding of the outcomes represents the different timings of the health effects. Abbreviations: w/=with; w/o=without; WHO=World Health Organization

## Discussion

### Summary

This article presents a consensus-building study using the Delphi methodology with the goal of co-developing a conceptual model of malaria during pregnancy with a diverse group of expert stakeholders. The result is a comprehensive disease policy model including ten maternal and ten child outcomes with four stratifiers. To our knowledge, it is the first formal attempt to co-develop a disease model of this kind either in the field of malaria or in a disease area predominantly prevalent in low and middle income countries.

The study has highlighted the complexity of the model required to depict appropriately the consequences of malaria during pregnancy to mothers and their offspring. Key contributors to the success of the study were the selection of the expert panel, thorough preparation of each stage as well as well as careful analysis and weighing-up of all responses. It was essential to be accurate with language, which sometimes had to evolve over various stages, while remaining accessible to a wide range of readers.

The process not only helped to develop the model to include relevant outcomes and relationships, but also improved the visual presentation and accessibility of the model, for example by adding symbols for the different timings of outcomes or appearance of arrows.

### Strength and limitations

This study has a number of strengths and limitations. The literature search conducted by the first author during the preparation stage was not a systematic review. Therefore some potential outcomes and relationships may have been missed out of the first draft of the model, however this was mitigated by the experts’ responses during the consultation rounds and consensus meetings. Experts for the Delphi panel were purposively selected to balance the experience, origin and focus area of work of panel members, however, the study may suffer from bias by omitting other experts with differing views.

The acceptance rate of experts was high (71%) with a 100% retention during the two consultation rounds; and 75% of panellists attended one of the two consensus meetings. The use of the Delphi methodology preserved the anonymity of experts and allowed panellists to respond freely without being influenced by other opinions or dominant personalities. The final stage of the study using online consultation meetings was more susceptible to the effects of group dynamics, however, this did not appear to be a problem with all experts engaging equally and respectfully with each other in both meetings.

This study focused on *P. falciparum* malaria. To apply the model to other plasmodium species such as *P.vivax*, *malariae* or *ovale* it would be necessary to review the model and consider inclusion of additional outcomes, relationships and stratifiers, informed by a literature search and expert consultation.

It may be a challenge to populate this comprehensive model for a cost-effectiveness study because of the range of outcomes and complexity of relationships. Nevertheless, this study has brought together experts from different fields and contexts to develop a model all could agree to.

### Areas for future research

During the study a number of areas requiring further research or development have emerged. The most commonly used outcome in cost-effectiveness analysis of global health interventions, the DALY, is a composite outcome combining mortality and morbidity, and in the case of malaria during pregnancy can combine both maternal and child outcomes into one measure. However, not all outcomes lend themselves equally well to calculating reliable DALY estimates and all of them rely heavily on assumptions made in the Global Burden of Disease studies (23). For example, estimating DALYs arising from “Long-term neurological and other sequelae” could potentially be difficult as long-term follow up data are lacking from malaria trials, requiring assumptions. Likewise, not all manifestations of severe malaria are equally associated with mortality or long-term morbidities, ultimately affecting the DALY. Placental malaria and asymptomatic parasitaemia were included in the model after the first consultation round, because for the experts, in particular epidemiologists, it is important to have these intermediate and often reported outcomes represented in the model for completeness and to depict important pathways.

Experts expressed very differing views regarding the inclusion of hypertension disorders of pregnancy, mostly shaped by different levels of awareness. The votes as well as the comments provided in the Delphi consultation indicated that experts working in lower endemicity settings were more aware of the link between malaria during pregnancy and hypertension disorder during pregnancy. The consensus meetings provided a useful platform to discuss these differences and the supporting evidence. Evidence from both Asia (24) and Africa points to an association, with a meta-analysis including four case-control studies from Africa suggesting that women experiencing malaria during pregnancy had 2.7 times higher odds of developing gestational hypertension disorder compared with those who did not (25). Ideally, the model should also differentiate hypertension disorder of pregnancy further into pre-eclampsia and eclampsia, but experts agreed that this level of detail will be difficult to find in the currently available data, but should be sought in the future. Experts commented that more effort should be made to collect data on hypertension disorder in pregnancy as blood pressure is always measured but often omitted from clinical trial databases. As above for “long-term neurological and other sequalae” the estimation of DALYs arising from hypertension disorder of pregnancy will require some assumptions on incidence and disability weights.

Some of the morbidities and outcomes can have lifelong consequences and be progressive. For example “neurocognitive and physical development impairment in <5” will likely impact the child for its entire life and influence its educational achievement and productivity. Likewise, for a women with severe malaria who develops severe anaemia and requires a blood transfusion there is a risk that the blood supply is contaminated, which then increases the risk of a wide range of other morbidities associated with other infectious diseases. While it will not be possible to quantify these future consequences for a cost-effectiveness analysis with currently available data, it is certainly important to create awareness of the potential long term health problems.

This study identified four important stratifiers: gravidity, transmission intensity, timing of exposure and HIV status. However, this does not preclude other variables from being important in certain analyses. Examples could be the sickle cell trait or the gender of the baby. At this point in time insufficient data are available to differentiate the consequences of the timing of exposure (e.g. first versus second, third trimester). HIV status also requires further disaggregation of the data such as the CD4 count or whether the woman is receiving antiretrovirals. Currently, cost-effectiveness models of chemoprevention for malaria during pregnancy will naturally stratify by HIV status as different prevention interventions are given to HIV negative and positive women.

Currently, the model does not include potential treatment or prevention interventions to ensure it is widely applicable for different purposes. Depending on the type of intervention study and context, further outcomes, such as side-effects, or effects on other diseases, such as HIV transmission from mother to child in HIV positive women, might need to be incorporated into the model. Studies using QALYs might also need to include patient’s health perception, perceived quality of life and future outlook into the model.

Finally, an important outcome of this study is to identify areas where data are scarce and share these with the research community, to raise awareness of the need for comparable outcome measures reported by trials.

A number of these points highlight the urgent need for more granular data, and experts felt that despite the complexity of the model it was important to create awareness of the wide range of outcomes that can be prevented by preventing pregnant women from being exposed to *P. falciparum*. More detailed data in the future should allow a move away from one size fits all models to models that are more adaptable and fluid.

## Conclusions

This study has demonstrated a more inclusive approach to developing disease policy models that are capable of assisting in the design of clinical trials (and other policy evaluations) and their associated health economic analysis. In so doing, we believe that this integrated approach should become the gold-standard for disease modelling designed to inform health policy in different countries and contexts. Co-development ensures wider perspectives are incorporated into the model than is usually possible for a single academic team, which should ensure the resulting model is more robust and fit for purpose. A robust conceptual modelling co-design approach will also help identify data gaps, ensuring these are not overlooked as the modelling proceeds to the implementation phase.

## Data Availability

Due to the small size of the Delphi panel, the detailed data collected during the Delphi consultations cannot be made available as individuals participating in the Delphi panel can be easily identified. However, a detailed analysis report, with all identifying information removed has been submitted for both rounds of Delphi consultations as an appendix.

## Contributors

All authors conceived the idea for the study. SF designed the study with input from KH and AB. SF conducted the Delphi consultations & meetings and subsequent data analysis. KH verified the data analysis. SF wrote the first draft of the manuscript. KH and AB critically reviewed and edited several drafts of the manuscript. All authors had full access to all the data in the study and had final responsibility for the decision to submit for publication.

## Acknowledgement

The authors would like to take this opportunity to acknowledge and thank the members of the Delphi Panel and Steering group for giving up their precious time so selflessly to share their invaluable wisdom with the study team.

**Delphi panel members (**in alphabetical order): Prof. Grant Dorsey (University of California San Francisco, United States of America), Prof. Kevin Kain (University of Toronto, UHN-Toronto General Hospital, Canada), Dr. Kassoum Kayentao (Malaria Research and Training Center-University of Sciences, Techniques, and Technologies of Bamako, Mali), Prof. Feiko ter Kuile (Liverpool School of Tropical Medicine, United Kingdom), Prof. Rose McGready (Shoklo Malaria Research Unit, Thailand), Dr. George Mtove (National Institute for Medical Research, Tanzania), Dr. Catherine Pitt (London School of Hygiene and Tropical Medicine, United Kingdom), Prof. Stephen Rogerson (University of Melbourne, Australia), Dr. Makoto Saito (WorldWide Antimalarial Resistance Network), Dr. Steve Taylor (Duke University, United States of America), Prof. Halidou Tinto (Institut de Recherche en Sciences de la Santé, Clinical Research Unit of Nanoro, Burkina Faso) and Dr. Holger Unger (Menzies School of Health Research, Darwin, Australia).

**Steering group members**: Prof. Kara Hanson (London School of Hygiene and Tropical Medicine), Prof. Andy Briggs (London School of Hygiene and Tropical Medicine), Prof. Feiko ter Kuile (Liverpool School of Tropical Medicine), Silke Fernandes (London School of Hygiene and Tropical Medicine)

## Overview of figures, tables and appendices

**Figures in appendix 1:**

- Figure S1: Conceptual model: Draft 2- Delphi consultation post round 1
- Figure S2: Conceptual model: Draft 3- Delphi consultation post round 2

**Appendices:**

- Appendix 1: Detailed methods and results
- Appendix 2: Questionnaire Delphi consultation round 1
- Appendix 3: Questionnaire Delphi consultation round 2
- Appendix 4: Summary report Delphi consultation round 1
- Appendix 5: Summary report Delphi consultation round 2

## Abbreviations

CEA: Cost effectiveness analysis
DALY: disability-adjusted life years

